# Heart Rate fractality disruption as a footprint of subthreshold depressive symptoms

**DOI:** 10.1101/2021.12.28.21268480

**Authors:** Piergiorgio Mandarano, Paolo Ossola, Pierluca Marazzi, Maria Carsillo, Stefano Rozzi, Davide Lazzeroni

**Author notes:** Corresponding author Piergiorgio Mandarano, School of Child and Adolescent Neuropsychiatry, Department of Clinical & Experimental Sciences, University of Brescia, Italy. These two authors contributed equally (Piergiorgio Mandarano, Paolo Ossola). These two authors contributed equally (Stefano Rozzi, Davide Lazzeroni).

## Abstract

**Introduction:** Psychopathology, and in particular depression, is a cardiovascular risk factor independent from co-occurring pathology. This link is traced back to the mind-heart-body connection, whose underlying mechanisms are, to date, not completely known. To study psychopathology in relation to the heart, it is necessary to observe the autonomic nervous system, which mediates among the parts of that connection. Its gold standard of evaluation is the study of heart rate variability (HRV).

**Objective:** To assess whether any association exists between the HRV parameters and sub-threshold depressive symptoms in a sample of healthy subjects

**Methods:** Two short-term HRV recordings (5 min - supine and sitting) were analysed in 77 healthy subjects. Here we adopted a three-fold approach to evaluate HRV: a set of scores belonging to the time domain (SDNN, pNN50, RMSSD); to the frequency domain (high, low, and very low frequencies) and a set of ‘nonlinear’ parameters. The PHQ-9 scale was used to detect depressive symptoms.

**Results:** Depressive symptoms were associated only with a parameter from the non-linear approach and specifically the long-term fluctuations of fractal dimensions (DFA-α2). This association remained significant even after controlling for age, gender, BMI, arterial hypertension, anti-hypertensive drugs, dyslipidaemia, and smoking habit. Moreover, the DFA-α2 was not affected by the baroreflex (postural change), unlike other autonomic markers.

**Conclusion:** In conclusion, fractal analysis of HRV (DFA-α2) allows to predict depressive symptoms below diagnostic threshold in healthy subjects regardless of their health status. DFA-α2 may be then considered as an imprint of subclinical depression on the heart rhythm.

## INTRODUCTION

Subthreshold depressive symptoms are commonly experienced in the general population, and they are not only a predictor of major depressive disorder but also of cardiovascular diseases. It is therefore crucial to develop an integrative understanding of this domain across multiple units of analysis from genes to neural circuits to behaviour (Woody & Gibb, 2015). Depression has been linked to an elevated risk of ischemic heart disease, myocardial infraction, and stroke in numerous meta-analyses (Gan et al., 2014; Van der Kooy et al., 2007; Woody & Gibb, 2015). Based on these findings, the American Heart Association (AHA) published a scientific statement in 2020 suggesting that depression is an independent risk factor for recurrent cardiovascular events in ACS survivors (Levine et al., 2021). The mechanisms underlying this mind-heart-body connection are not fully understood however the autonomic nervous system seems to play a crucial role in mediating this relationship(Levine et al., 2021). Supporting this hypothesis, genetic background of subjects with depressive symptoms explains most of the variance of the HRV, suggesting that a common neurobiological dysregulation links depressive symptoms and the heart (De Jonge et al., 2007; Su et al., 2010; Vaccarino et al., 2008).

The gold standard analysis of the autonomic nervous system, indicated by the GRAPH guidelines (Quintana et al., 2016), is the study of heart rate variability (HRV). The variability of the heart rate is studied starting from the electrocardiographic data considering the R-R intervals’ distribution in a chosen timeframe, named tachogram. The tachogram is studied through three groupings of parameters according to temporal criteria, decomposition in frequency bands and nonlinear calculations (***Table 1***) (Shaffer & Ginsberg, 2017). A high variability in the tachogram is a sign of a healthy response of the parasympathetic system, which, in cardiac patients is associated with a better survival (Malik et al., 1996; Shaffer & Ginsberg, 2017). The main problem when exploring the HRV through the classic parameters is their ecological validity. In fact, most of these parameters mirror the sympathetic tone that is affected, among the others, by posture through the baro-reflex (Malik et al., 1996).

**Table 1.** HRV measures

| Parameter | Unit | Description |
| --- | --- | --- |
| **HRV time-domain measures** |  |  |
| SDNN | ms | Standard deviation of NN intervals |
| pNN50 | % | Percentage of successive RR intervals that differ by more than 50ms |
| RMSSD | ms | Root mean square of successive RR interval differences |
| **HRV frequency-domain measures.** |  |  |
| VLF | Hz | Very low-frequency band (0.0033–0.04Hz) |
| LF | Hz | Low-frequency band (0.04–0.15Hz) |
| HF | Hz | High-frequency band (0.15–0.4Hz) |
| LF/HF |  | Ratio of LF-to-HF power |
| **HRV non-linear measures.** |  |  |
| Sample Entropy |  | Sample entropy, which measures the regularity and complexity of a time series |
| DFA $\alpha_1$ | | Detrended fluctuation analysis, which describes short-term fluctuations |
| DFA $\alpha_2$ | | Detrended fluctuation analysis, which describes long-term fluctuations |

*Legend: IBI, Interbeat interval; time interval between successive heartbeats; NN intervals, interbeat intervals from which artifacts have been removed; RR intervals, interbeat intervals between all successive heartbeats.*

A higher heart rate variability reflects, also, a better physiological capacity of flexible emotion regulation in response to stress (Thayer et al., 2012). This pathogenic mechanism parallels the behavioural one, according to which depressive symptoms are often the results of a maladaptive response to triggering events (Hammen, 2005). A reduced HRV has already been shown to be an independent predictor of clinical depression (Westhoff-Bleck et al., 2021) and a low vagal tone has been proposed to play an etiologic role in early-stage depressive symptoms development (Jandackova et al., 2016). In fact, the HRV predictive power for depression is comparable to that of other tools like the PHQ-9 (Economides et al., 2020; Pizzoli et al., 2021), the most accurate DSM-V based screening questionnaire (Kroenke et al., 2010).

When exploring the association between heart rate variability and mood symptoms, the use of non-linear calculations seems crucial to the description of mind-heart-body interactions - in term of affect and cognition - that are not captured by linear analyses of time and frequency (Jung et al., 2019; Pham et al., 2021).

A few papers evaluated the association between non-linear parameters and depression (Blasco-Lafarga et al., 2010; Byun et al., 2019; Fiskum et al., 2018; González et al., 2013; Perkiömäki, 2011; Vigo et al., 2004) but often they included patients with a long-lasting history of cardiac or depressive illness, medicated, or with other comorbidities that might hinder the true association between depressive symptoms and the HRV parameters.

Thus, the present study has two aims. First, evaluate the independency of each HRV parameter from the baro-reflex comparing each measurement when supine and seated. Second, assess whether any association exists between the HRV parameters and sub-threshold depressive symptoms in a sample of healthy subjects.

## MATERIALS

The Don Gnocchi Foundation ethics committee approved the study. After the study was fully explained all the participants signed an informed consent.

### STUDY DESIGN AND POPULATION

We recruited n=77 healthy subjects from the Cardiovascular Prevention Unit of the Don Gnocchi Foundation in Parma between January 2017 and December 2019

This was part of a larger sample of 1.130 subjects as part of a 5-year prospective registry of patients who underwent cardiovascular evaluation. Out of the original sample we first excluded those with an invalid ECG signal (n=376). Out of the remaining 754 patients, we excluded n=72 subjects who did not complete the PHQ-9 questionnaire and n=94 who did not completed the PSS-10. Two-hundred-seventy-nine subjects also had missing data on some socio-demographic variables resulting in a sample of n=309 subjects. After this initial selection, following the GRAPH guidelines (Quintana et al., 2016), we also excluded the patients: (A) older than 65 (n=467); (B) with a history of major cardio-vascular event (n=54) defined as a previous major cardiovascular event (myocardial infraction, chronic coronary syndrome or stroke) or other cardiovascular disease such as chronic heart failure, cardiomyopathies and cardiac valve diseases) (Quintana et al., 2016); (C) with diagnosis of diabetes (n=30) defined as fasting glycaemia 126 mg/dL (≥7.0 mmol/L) (Cosentino et al., 2020); (D) taking beta-blocker medications (n=21); and (E) with a diagnosis of Major Depressive Episode according DSM-5 (APA, 2013) (n=21).

The study flowchart is depicted in **Figure 1**.

**Figure 1.**
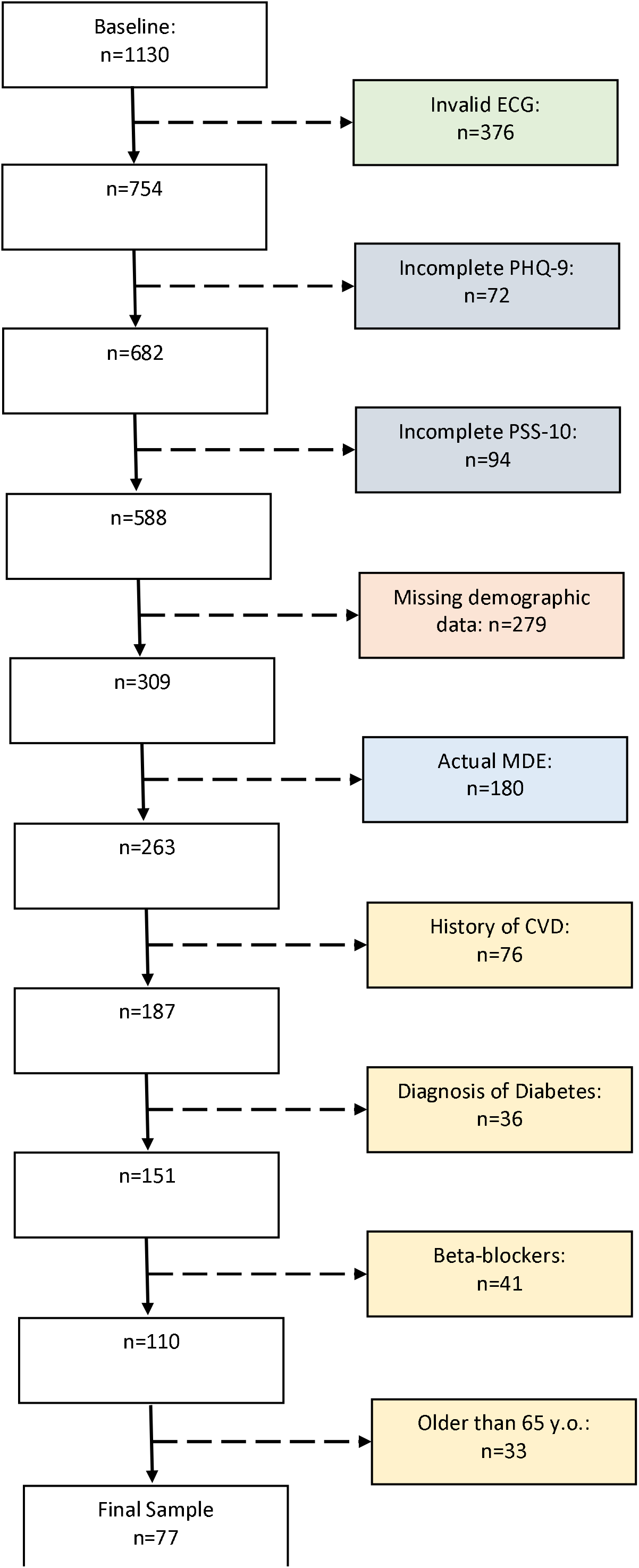
Study Flowchart **Note**. PHQ-9= Patient Health Questionnaire-9 is a self-reported questionnaire assessing depressive symtpoms; PSS-10 = Perceived Stress Scale-10 is a self-report questionnaire measuring the subjective stress; MDE=Major Depressive Episode; CVD=cardiovascular disease.

The first part of the experiment consisted in a semi-structured interview aimed at collecting socio-demographic and cardiovascular risk information. These were age, gender, BMI, cigarette smoke, dyslipidaemia, blood hypertension, and antihypertensive drugs. Based on antihypertensive medications subjects were divided into with and without medications (codes as 1 and 0 respectively). Dyslipidaemia was defined as at least one among total cholesterol ≥200 mg/dL (5.172 mmol/L); LDL≥130 mg/dL (3.3618 mmol/L); Triglycerides >150 mg/dL (1.6935 mmol/L); or HDL-C, <35 mg/dL (0.9051 mmol/L). Hypertension was defined as having systolic blood pressure greater than 140 mmHg or a diastolic blood pressure greater than 90 mmHg or anti-hypertensive drugs.

Then each subject filled two self-reported questionnaires. Specifically, one aimed at evaluating the depressive symptoms (PHQ-9) and one aimed at evaluating the previous exposure to stressful events (PSS-10).

The PHQ-9 (Patient Health Questionnaire-9) is a self-reported questionnaire of 9 items on a Likert scale (from 0 to 3) based on the DSM-5 criteria for a Major Depressive Episode (Kroenke et al., 2001). A score above 10 was found to be 88% accurate in diagnosing depression and hence it was the suggested cut-off (Arroll et al., 2010). This tool has been developed in the first place to diagnose major depressive episodes in primary care. Previous studies adopted the PHQ as a dimensional construct to assess depressive symptoms (Chen et al., 2006).

The PSS-10 (Perceived Stress Scale-10) is self-administered questionnaire of 10 items on a 5-points Likert scale aimed at gauging the perceived stress in the previous month (Cohen & Williamson, 1988; Mondo et al., 2021). The shorter version with 10 items (Lee, 2012) showed a good internal consistency.

In a second part of the experiment on the same day, ECG recording was collected in a paradigm complying to the Guidelines for Reporting Articles on Psychiatry and Heart rate variability (GRAPH) (Quintana et al., 2016). In details, patient have been recorded in a quiet environment at 24°C, in the morning, with uncontrolled breathing rate, in supine position at rest. Inter-beat-intervals (IBI) were extracted from an ECG track recorded over a 15-min period with the Nexfin device (Edwards Lifesciences) at 200 Hz. Following this acquisition, the patients were required to seat, and the ECG was recorded in this position, in the same conditions described above. In both cases, to catch the 5-min with less artefact a 5-min sample was chosen from the cleanest ECG derivation of the 6 recorded. The acclimation period and all the artefacts have been manually removed by an expert cardiologist (DL).

## ECG ANALYSES

To evaluate the ECG, we adopted a three-fold approach. Specifically, we computed three groups of parameters: (a) the Time domain, defined by SDNN, pNN50, and RMSSD; (b) the Frequency domain that computes high (HF), low (LF) and very low (VLF) frequencies; and (c) the complexity ‘nonlinear’ analyses, namely Sample Entropy and two Detrended fluctuation analysis (DFA-α1; DFA-α2) (Quintana et al., 2016; Shaffer & Ginsberg, 2017)

The degree of variability of the inter-beat interval (IBI), which is the time interval between successive heartbeats, is quantified using time-domain indices of HRV. Three Time domain parameters are computed from the tachogram that depict the RR intervals over time (ms). The SDNN is the standard deviation of the normal R-R intervals (N-N intervals). SDNN is the “gold standard” for medical stratification of cardiac mortality risk where greater SDNN means greater HRV and hence lower risk (Kleiger et al., 1987). The RMSSD is the square root of the mean squared differences of successive N-N intervals, and usually offers a more accurate estimation of the parasympathetic function because it is less influenced by respiratory sinus arrhythmia (Penttilä et al., 2001).

The pNN50 is the percentage of successive RR intervals that differ by more than 50⍰ms. pNN50 has been linked to parasympathetic activity (Otzenberger et al., 1998).

From the tachogram we derived the corresponding power spectral density (PSD) that defines the range of possible frequencies of the tachogram in a finite time interval. The range of frequencies is then divided into three main components: very low (VLF) between 0.0033–0.04⍰Hz, low (LF) between 0.04–0.15⍰Hz and high frequencies (HF) between 0.15–0.4⍰Hz. The VLF band reflects the activation of multiple levels of feedback and feed-forward loops in the heart’s intrinsic cardiac neural system, as well as between the heart, the extrinsic cardiac ganglia, and the spinal column (Kember et al., 2001; Taylor et al., 1998). The LF shows how the breath rate influence the efferent vagal fibres (Di Nardo et al., 1993). The HF has been considered the purest marker among HRV to measures the vagal tone (parasympathetic activity) (Egizio et al., 2011).

A third approach for the analysis of the tachogram is the non-linear dynamics. Doing so we can quantify the amount of regularity and the unpredictability of fluctuations over time-series data. Entropy measures the degree of randomness in the cardiovascular system quantifying the unpredictability of fluctuations in the RR time series. A tachogram characterised by low entropy would hence show low variability being ordered and repetitive. ON the contrary, healthy subjects would display a greater variability with high entropy in the tachogram. Sample entropy is a regularity statistic that.

Based only on this parameter we can potentially differentiate between a healthy ECG with a high degree of regularity and an irregular and unpredictable one in which the RR intervals are randomly spaced. Literature seems to agree that Sample Entropy reflects how much the variations in consecutive RR intervals are unpredictable (Shaffer & Ginsberg, 2017).

Another method to evaluate regularity is the self-affinity. Self-affinity defines how much a given time-series is exactly or approximately like a part of itself. As for the fractals, this means that the whole has the same shape as one or more of the parts. Detrended fluctuation analysis (DFA) introduced by Peng et al. (1995) quantifies the fractal scaling properties of time series and is a method for determining the statistical self-affinity of a signal. This analysis can be done both on short-term intervals (shorter than 30 s) (DFA-α1) and long-term intervals (>30 s) (DFA-α2). The scaling exponent indicates the strength of the correlations with previous values in the time series. A value of 1 would suggest a fractal-like process in which the fluctuations are generated by multiple feedback regulations. Values close to 0.5 are indicative of random dynamics; values near 1.5, on the other hand, describe a highly correlated pattern

The DFA-α1 is a baroreflex estimate, and strong variations of this parameter are linked to increased cardiovascular, neurological, and mental morbidity and mortality. The DFA-α2 has been a part of the panel since the 1980s, and it has only shown connections with non-pathological states like circadian rhythm and age (Beckers et al., 2006; Iyengar et al., 1996; A. Voss et al., 2012). Decreased values of DFA-α1 and DFA-α2 correspond to worse outcome after myocardial infarction (Tapanainen et al., 2002) or worse quality of life (Seifert et al., 2014).

The explanation of the acronyms and the units of these metrics are shown in **Table 1**.

## STATISTICAL ANALYSES

Because of the skewed distribution of the ECG parameters (n=754) and of the depressive symptoms (n=682) we applied a Rank-based inverse transformation (RIN) transformation (Bishara & Hittner, 2015). RIN transformation, as in the Spearman approach, involves converting the data into ranks. The ranks are then converted into probabilities that, with inverse cumulative normal function, are transformed into an approximately normal shape.

The normality of each variable (natural and RIN-transformed) has then been tested with the Shapiro-Wilk (W) test. The more the W approaches to 1, the more the distribution is normal (**Table S1** and **Figure S1**).

According to the primary objective we then explored the association between the depressive symptoms (outcome) and the ECG parameters (predictors) through bivariate Pearson correlations in the final sample (n=77). Because of multiple correlations between the PHQ and the ECG parameters (n=10) we adjusted the p-value with a Bonferroni correction so that in these exploratory analyses the p-value for a threshold of significance was set at 0.005.

We then looked at whether the afore-mentioned association was influenced by any registered condition. We then build a model (multiple linear regression) with depressive symptoms as our outcome and the parameter as the predictor of interest controlling for socio-demographic and clinical variables that might affect this association. To avoid multicollinearity, we also reported the Variation Inflation Factor (VIF). A value of VIF greater than 4 suggest a multicollinearity problem.

Final model for healthy subjects:

PHQ-9 (RIN-transformed) = b0 + b1*parameter (RIN-transformed) + b2*age + b3*gender + b4*BMI + b5*dyslipidaemia + b6*hypertension + b7*smoking habit + b8*Anti-hypertensive therapy (a priori without beta-blockers) + b9*PSS-10

Where gender is coded as 0=female, 1=male; BMI=body mass index; dyslipidaemia coded as 0=normal, 1= yes; Hypertension coded as 0=no; 1=yes; Smoking Habit coded as 0=no, 1=yes; Anti-hypertensive therapy (a priori without beta-blockers) coded as 0=no, 1=yes; PSS-10= Perceived Stress Scale.

Lastly, to further test the ecological validity of the parameters associated with depressive symptoms, we explored whether these varied from supine to seated through Pearson correlation and paired t-test. This because a change in the parameters from supine to seated would suggest a significant effect of the baro-reflex on the parameter.

## RESULTS

The sample descriptive is summarised in **Table 2**.

**Table 2.** Sample (n=77) descriptive

| **Table 2. Sample (n=77) descriptive** |  |
| --- | --- |
| Age | 51.09 (9.53) |
| Gender (Male) (n, %) | 42 (54.5) |
| BMI | 26.58 (4.91) |
| Dyslipidaemia (n, %) | 53 (68.8) |
| Hypertension (n, %) | 59 (76.6) |
| Smoking Habit (n, %) | 31 (40.3) |
| Anti-hypertensive therapy (n, %) | 26 (33.8) |
| PHQ-9 | 4.58 (2.54) |
| PSS-10 | 17.70 (5.79) |

**Note.** The values refer to the mean and standard deviation if not otherwise specified. BMI: Body Mass Index (Kg/m<sup>3</sup>). CVD: cardio-vascular diseases.

When exploring the association through Pearson correlations between the HRV parameters and the depressive symptoms only the DFA-α2 reached the statistically significant threshold adjusting for multiple comparisons (**Table 3, Figure 2**)

**Table 3.** Association between depressive symptoms at PHQ and ECG parameters RIN transformed

|  | **Pearson's r (p-value)** |
| --- | --- |
| SDNN | 0.027 (0.813) |
| pNN50 | 0.183 (0.110) |
| RMSSD | 0.150 (0.193) |
| VLF | -0.101 (0.382) |
| LF | 0.004 (0.971) |
| HF | 0.204 (0.075) |
| LF/HF | -0.272 (0.017) |
| Sample Entropy | 0.128 (0.267) |
| DFA-$\alpha_1$ | -0.263 (0.021) |
| DFA-$\alpha_2$ | -0.326 (0.004) |

**Note.** pNN50 percentage of successive RR intervals that differ by more than 50ms, RMSSD square root of the mean squared differences between successive RR intervals, VLF very low-frequency power, LF low-frequency power, HF high-frequency power, LF/HF LF/HF ratio of low-frequency and high-frequency power, DFA- $\alpha_1$ detrended fluctuation analysis short-term parameter, DFA- $\alpha_2$ detrended fluctuation analysis long-term parameter.

**Figure 2.**
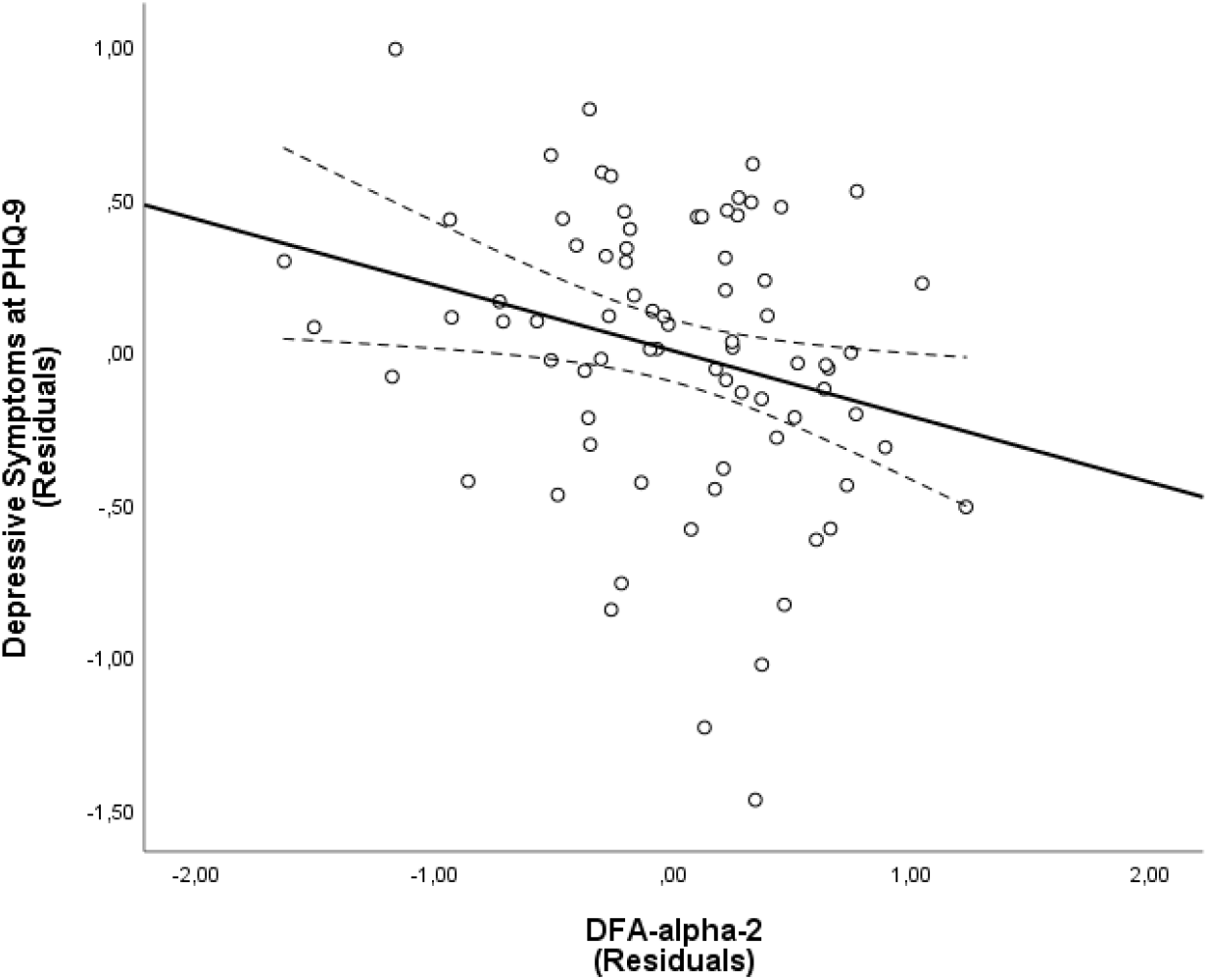
Association between DFA-α2 and PHQ. Note. Higher DFA-α2 values are associated with more severe depressive symptoms (higher PHQ-9 score). The Y-axis represents the residuals in predicting the depressive symptoms from all the independent variables included as a control in the model for Healthy Subjects. These are the subjects’ perceived stress, age, gender, BMI, dyslipidaemia, hypertension, smoke, hypertensive. The X-axis represents the residuals from predicting PHQ from the same independent variables, that is the full model without the DFA-α2. Dotted lines represent the 95% confidence interval.

The negative association between DFA-α2 and depressive symptoms remained significant even when controlling for other socio demographic variables (beta=-0.227, p=0.022) or previous stressful events at PSS-10 (beta=-0.239; p=0.028) (**Table S2**). This means that higher the value at detrended long-term fluctuation analysis (DFA-α2) the worse the depressive symptoms according to the PHQ-9 scale.

In our sample DFA-α2 and PHQ-9 were not associated with age, BMI nor varied as a function of gender, smoking habit, dyslipidaemia, hypertension, or hypertensive medication.

As expected for a parameter gauging the parasympathetic system the difference between the two was not significant (t=-0.491; p=0.625) (**Table S3**). This suggests that the baroreflex does not significantly affect this parameter that could represent a good ecological measurement.

The DFA-α2 supine was the only variable that did not correlate with its corresponding value when seated (r=.194, p=0.091). This correlation however was moderated by depressive symptoms at PHQ-9 (interaction: B=-0.1167; 95%CI=-0.2024, -0.0311; SE=0.043; t=-2.716; p=0.008) so that the association between DFA-α2 when supine and seated was true and positive only at low (P=0.0021) and average (p=0.0251) levels of depressive symptoms.

## DISCUSSION

Our findings show that sub-threshold depressive symptoms are associated with the long-range fractal fluctuations of the heart rate. In particular, healthy subjects with greater depressive symptoms had more randomness in the tachogram when considering intervals longer than 30 seconds. This means that the more depressed a subject is, the lower is the fractal tendency over long-term intervals in the heart-rhythm. The autonomic underpinning of this correlation is now largely unknown, we hypothesize that it reflects an excessive parasympathetic activation. This is in line with evidence indicating that an increase in parasympathetic influence decreases exponent alpha values and indicates more random autonomic behaviour (Shaffer & Ginsberg, 2017).

This finding is important for understanding not only the relation between autonomic nervous system and hearth, but also between depressive symptoms and the autonomic system. We can hypothesise that a greater randomness in autonomic activation and higher depressive symptoms are both a measure of ineffective self-regulation. Accordingly, a subject with low self-regulatory abilities when facing a stressor would be more likely to develop depressive symptoms on a cognitive-affective point of view and to increase the randomness of their HRV on a neuro-physiological level. Specifically, external stressors would immediately impact on the sympathetic branch of the autonomous nervous system increasing the stiffness of short-term fractals that are classically associated with anxiety symptoms. More on the long run it is possible that vulnerable subjects (e.g., low in effortful control) would develop depressive symptoms associated with an increased parasympathetic tone that explains the increase randomness of long-term fractals. Given the cross-sectional design of the study we can’t establish what come first, but we can hypothesise that a loss of the fractal properties of the tachogram is the way depression exert its negative effect on cardiac outcome (Ossola et al., 2018).

Our hypothesis seems supported by the fact that DFA-α2 is also a measure of top-down control, which is crucial in emotion regulation (Wager et al., 2008). A previous study demonstrated that DFA-α2, but not DFA-α1 or sample entropy, was greater when the subjects faced harder trials in a task tapping specifically the prefrontal functions (Mukherjee et al., 2011).

Previous studies found that the long-term fractals dimensions are lower in male than in female (Beckers et al., 2006; Andreas Voss et al., 2015). This is in line with findings in emotion regulation that show less effective strategies in men (Nolen-Hoeksema, 2012) that are generally associated with more depressive symptoms (Nolen-Hoeksema & Aldao, 2011).

Altogether these findings point to long-range fractal fluctuations as a neurophysiological mechanism mirroring the self-regulatory abilities. Note, however, that, among the cortical regions, the prefrontal cortex is characterized by the widest paths of connections and participate to several distinct cognitive and emotional functions. Accordingly, for a physiologically grounded understanding of this relation, more specific investigations are needed to further support our hypothesis and unveil the underpinning functional mechanism

Literature has often evaluated overt depression through the study of HRV in the time and frequency domain. Results consistently point at an association between depressive symptoms and those parameters strongly related to DFA-α2 such as the ratio between VLF and the sum of LF and VLF (Blood et al., 2015; Borrione et al., 2018; Carney et al., 2005; Francis et al., 2002; Jain et al., 2014; Kemp et al., 2010; Paniccia et al., 2017).

Only one previous study found an association between DFA-α2 and depressive symptoms (Kojima et al., 2008) but in the opposite direction to ours. The association however was not linear and driven mostly by higher depressive scores in the last quintile of DFA-α2 with no differences in depressive symptoms among the other quintiles. It is possible that their results are due to the selected sample. The authors, in fact, recruited a group of chronic haemodialysis patients often with diabetes and beta-blockers that can alter the HRV parameters.

Previous studies found an association between depression and DFA-α1, but results are mixed. Some authors found a negative association with depression (Kojima et al., 2008), with lower levels of DFA-α1 in subjects with internalising psychopathology (Fiskum et al., 2018) and a higher rate of subjects with DFA-α1 lower than 1 in depressed subjects (Kop et al., 2010). Other, instead, found a positive association between depressive symptoms and DFA-α1 (Vigo et al., 2019; Vigo et al., 2004) and higher values of DFA-α1 in depressed subjects (Schultz et al., 2010).

Again, this could be due to patients with a recent unstable angina pectoris or acute myocardial infarction (Vigo et al., 2004) and clinically depressed subjects (Kwon et al., 2019). Again, all these were exclusion criteria from our final sample. We emphasize that our association carefully controls for an individual’s variable that are known to affect the heart rate variability such as stress perception, age, gender, BMI, smoking habit, dyslipidaemia and hypertension or anti-hypertensive medication. Notably, because also circadian rhythm seems to affect the DFA-α2 (Beckers et al., 2006) all the recording has been done at approximately the same daytime in a stable environment.

Importantly, our results demonstrate that the DFA-α2 parameter that reflects this self-affinity over long-term intervals, was not significantly affected by the baro-reflex (Uhlig et al., 2020). Studying the effect of activating the baroreflex, by comparing the two recordings in different postures, we note that purely parasympathetic markers, such as DFA-α2 remain constant. This can allow us to hypothesize that the predictivity of the DFA-α2 is independent from the postural change and that the subclinical depression acts on the heart through parasympathetic fibres. Interestingly DFA-α2 was the only parameters among those explored that did not correlated between the supine and the seated position. The association between DFA-A2 when supine and seated was moderated by depressive symptoms so that the more a depressed a subject is, the less correlated are the two measurements. The baro-reflex sensitivity is a measure of the functioning of a reflex loop involving pressure-sensitive nerves (i.e., baroreceptors) mainly in the carotid arteries and the aorta. High basal sympathetic activation, a condition frequently found in depressed (Broadley et al., 2005; Davydov et al., 2007), leads to a decline in the effective baroreflex sequences. It is possible that a sympathetic predominance, that is associated with DFA-α2 values greater than 1, explains the lack of association between the two values.

## CONCLUSIONS

Fractal analysis of HRV, evaluated as DFA-α2, can predict depressive symptoms below diagnostic threshold in the healthy subject, regardless of comorbidities and autonomic condition of the subject, thereby indicating a background radiation, a “footprint” that depressive symptoms leave on the heart rate. Beside the theoretical implications previously discussed, we believe that our study also has possible clinical implications in prevention and therapy follow-up.

In fact, it is known that often major depression is anticipated by a phase of subclinical depression (Ossola et al., 2015). There is evidence that treatment response rates are higher when treatment is started at the onset of sub-syndromic symptoms but at that time depression is more difficult to diagnose as patients are more prone to report physical symptoms rather than emotional ones (Simon et al., 2008). In this sense a screening tool such as DFA-α2 from the electrocardiogram would help in identifying patients at risk of developing a full-blown episode. That would exponentially enlarge the screening potential with an optimal cost-effectives, allowing not only to make prevention campaigns for depression but also for the resulting cardiovascular disease.

A second possible clinical implication of the present study is represented by the potential use of DFA-α2 in clinical practice as an autonomic marker in the follow-up of patients with sub-threshold and clinical depression. Further longitudinal studies evaluating HRV parameters before the onset of depressive symptoms and during the course of illness might help in disentangling this association.

We have called DFA-α2 the “footprint of depression”, but this does not exclude that the relationship between the two is not exclusive, since no research has yet been carried out to correlate this specific parameter to other psychopathologies (Alvares et al., 2016). Further studies should evaluate the validity of DFA-A2 in ecological settings such as Holter monitoring and test its specificity and predictive ability prospectively on depressive symptoms.

## Data Availability

All data produced in the present study are available upon reasonable request to the authors

## SUPPLEMENTARY

**Table S1.** Normality Distribution and RIN transformation in the whole sample (n=754)

|  | Natural | RIN-transformed |
| --- | --- | --- |
| SDNN | 0.860 (p<0.001) | 0.999 (p=0.999) |
| pNN50 | 0.644 (p<0.001) | 0.964 (p<0.001) |
| RMSSD | 0.850 (p<0.001) | 0.999 (p=0.999) |
| VLF | 0.476 (p<0.001) | 0.999 (p=0.999) |
| LF | 0.600 (p<0.001) | 0.999 (p=0.999) |
| HF | 0.415 (p<0.001) | 0.999 (p=0.999) |
| LF/HF | 0.789 (p<0.001) | 0.999 (p=0.999) |
| Sample Entropy | 0.981 (p<0.001) | 0.999 (p=0.999) |
| DFA-α1 | 0.986 (p<0.001) | 0.999 (p=0.999) |
| DFA-α2 | 0.993 (p=0.001) | 0.999 (p=0.999) |
| PHQ-9 | 0.908 (p<0.001) | 0.993 (p<0.001) |

**Note.** Here reported the Shapiro-Wilk test (W) and the p-value in parenthesis for each variable both as computed and after Ranked-base Inverse (RIN) transformation. Only n=682 subjects completed the PHQ.

**Figure S1.**
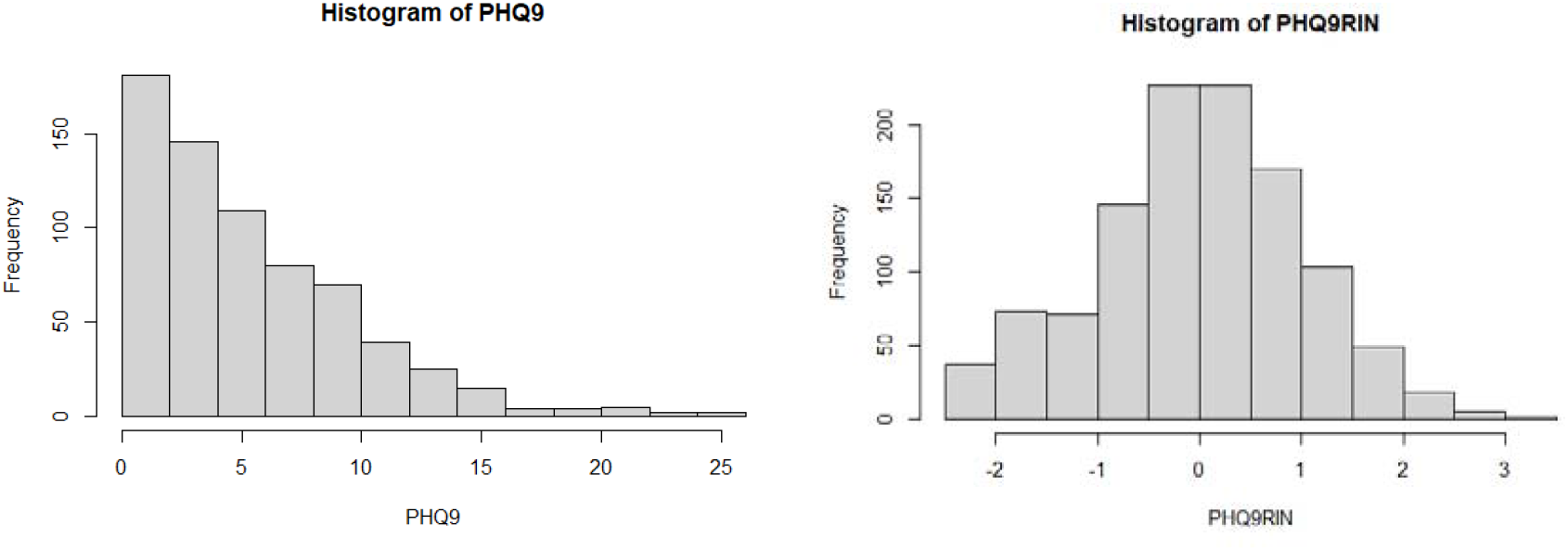
Example of distribution of PHQ before and after RIN transformation (n=682)

**Table S2.**
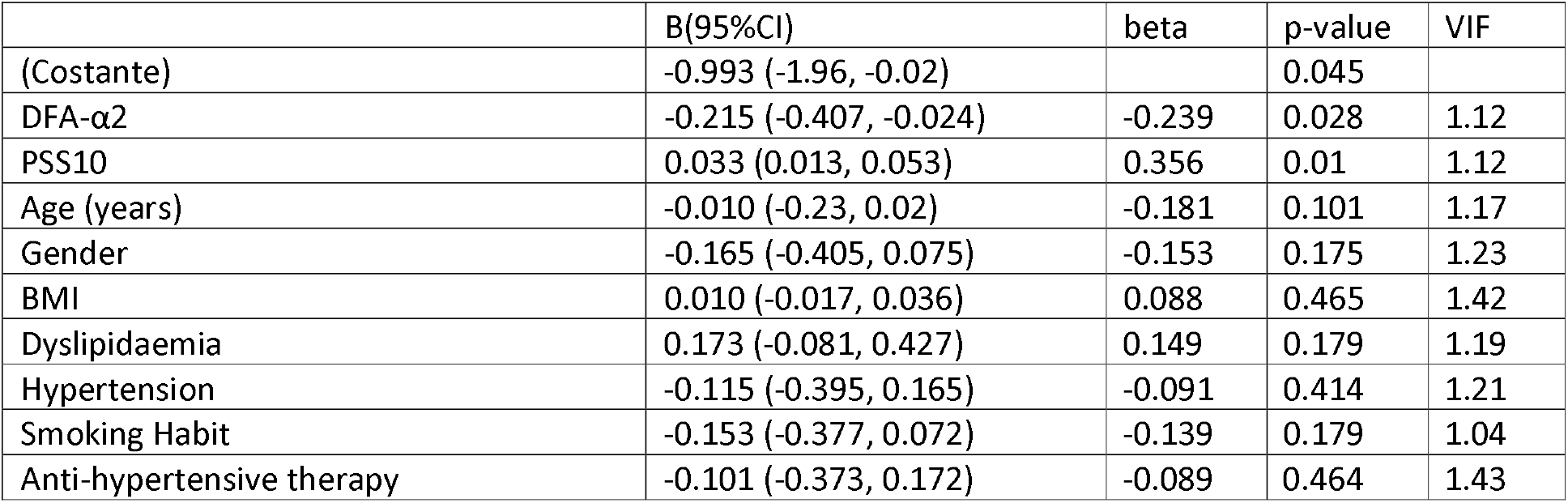
Association between DFA-α2 (RIN transformed) and PHQ-9 (RIN-transformed) controlling for confounding variable in a multiple linear regression (n=77)

**Table S3.** Change of HRV parameters after positional change.

|  | Supine (SD) | Seated (SD) | correlation | t | p-value |
| --- | --- | --- | --- | --- | --- |
| SDNN | 40.83 (19.52) | 43.28 (19.84) | 0.533 | -0.590 | 0.557 |
| pNN50 | 0.076 (0.11) | 0.055 (0.083) | 0.722 | -0.512 | 0.610 |
| RMSSD | 26.53 (15.59) | 24.71 (14.29) | 0.704 | -0.728 | 0.469 |
| VLF | 829.89(1090.86) | 944.01 (992.35) | 0.397 | -0.259 | 0.796 |
| LF | 555.34 (640.69) | 697.01 (872.71) | 0.564 | -0.290 | 0.773 |
| HF | 238.10 (274.08) | 214.98 (242.01) | 0.756 | -0.459 | 0.648 |
| LF/HF | 3.08 (1.927) | 4.55 (4.06) | 0.575 | -0.118 | 0.906 |
| Sample Entropy | 1.43 (0.281) | 1.30 (0.27) | 0.379 | 2.058 | 0.043 |
| DFA-α1 | 1.18 (0.224) | 1.28 (0.24) | 0.452 | -1.319 | 0.191 |
| DFA-α2 | 0.94 (0.163) | 0.94 (0.17) | 0.194 | -0.514 | 0.609 |

**Note.** The mean and standard deviation are reported for the natural value. The paired t-test and the spearman correlations instead are computed with the RIN transformed values. All the correlation are significant at a $p < 0.001$ level except DFA-2 ( $p = 0.091$ ).

**Figure S2.**
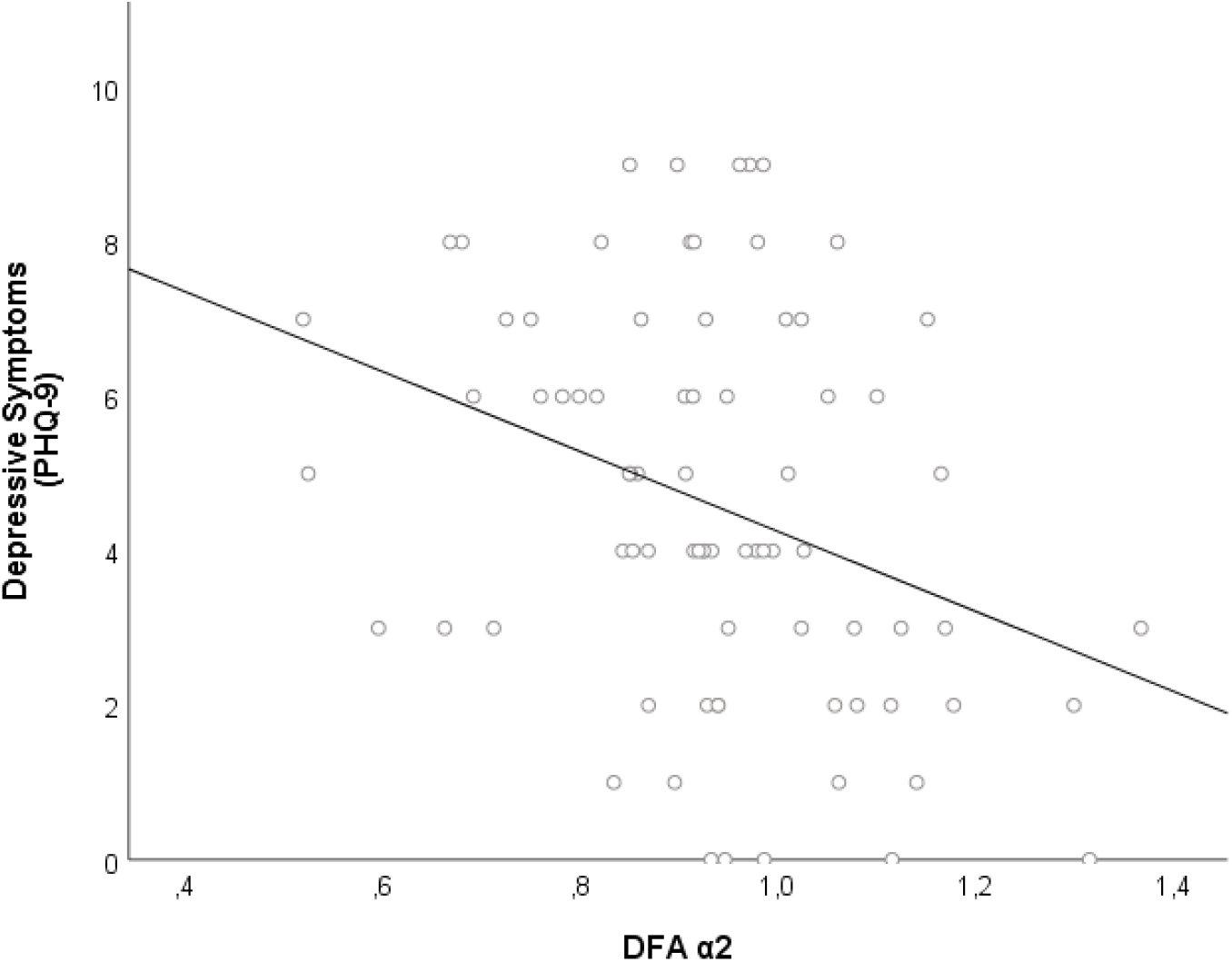
Association between depressive symptoms and DFA-2 **Note**. Negative correlation between DFA-2 and depressive symptoms at PHQ in n=77 subjects (r=-0.333; p=0.003). On the X axis the actual values of DFA-2. DFA-2 values around 1 indicate autonomic nervous system performance that is fractal and robust. Excessive parasympathetic effect lowers exponent alpha values, signaling more erratic autonomic behavior. Sympathetic predominance, on the other hand, results in alpha values larger than 1, indicating stiffer, more linked autonomic function. A deviation from the exponent alpha-1 value to either side indicates that the ANS’s complexity has deteriorated. On the Y-axis the actual values of PHQ-9.

